# The potential impact of microRNA-related functional polymorphisms in the pathogenesis of coronary heart disease

**DOI:** 10.1101/2023.10.09.23296433

**Authors:** Taqweem Ul Haq, Yasir Ali, Sami Ur Rehman, Sajjad Ali, Yangchao Chen, Fazal Jalil, Aftab Ali Shah

**Affiliations:** Department of Biotechnology, University of Malakand, Khyber Pakhtunkhwa, Pakistan; Department of Biotechnology, Abdul Wali Khan University Mardan, Khyber Pakhtunkhwa, Pakistan; School of Biomedical Sciences, Chinese University of Hong Kong, NT, Hong Kong

**Keywords:** microRNA, single nucleotide polymorphism, coronary heart disease, TaqMan assay

## Abstract

MicroRNAs (miRNA) are important post-transcriptional gene regulators. Various populations have experienced a marked rise in the risk of coronary heart disease (CHD) due to multiple miRNA variations. The current case-control study (150 cases and 150 healthy controls) was designed to determine the potential role of five miRNA functional variants (rs2292832, rs3746444, rs11614913, rs1044165, and rs767649) as risk factors for CHD in the Pakistani population using TaqMan Real-time PCR Assay. It was observed that the single nucleotide polymorphism (SNP) rs3746444 was significantly associated with the risk of CHD using the co-dominant model [χ2 =79.51; P = 0.0001], dominant model (GG vs AA+AG) [OR = 9.333 (5.180-16.82); P = 0.0001], heterozygous model (AG vs AA+GG) [OR = 0.1241 (0.065-0.234); P = 0.0001] and additive model [A vs G; OR = 0.3440 (0.2468-0.4795); P = 0.0001] respectively. Furthermore, rs11614913 was also linked with CHD when analyzed using a co-dominant model [χ2 =16.24; P = 0.0003], dominant model (CC vs CT+TT) [OR = 1.918 (1.210-3.042); P = 0.0075], recessive model (TT vs CT+CC) [OR = 0.2754 (0.1369-0.5540); P = 0.0002], and additive model [OR = 2.033 (1.445-2.861); P = 0.0001]. It was also found that rs767649 is connected to CHD using a co-dominant model [χ2 =114.9; P = 0.0001], dominant model (AA vs AT+TT) [OR = 7.851 (3.554-17.34); P = 0.0001], recessive model (TT vs AT+AA) [OR = 0.04956 (0.026-0.092); P = 0.0001], heterozygous model (AT vs AA+TT) [OR = 4.495 (2.737-7.382); P = 0.0001], and inheritance additive model [A vs T; OR=7.154 (4.902-10.44); P = 0.0001] respectively. The SNP rs1044165 revealed a strong correlation with CHD using the heterozygous inheritance model (AG vs GG+AA) [OR = 0.3442 (0.1308-0.9055); P = 0.0276]. No statistically significant association (P □ 0.05) of rs2292832 SNP with CHD was found using all five inheritance models.

## Introduction

According to the World Health Organization (WHO), coronary heart disease (CHD) is one of the leading causes of illness and mortality globally. [1]. Environmental and genetic factors have been implicated in the pathophysiology of CHD [2]. Environmental factors include lack of exercise, an imbalanced diet, a sedentary lifestyle, low socioeconomic status, hypertension, diabetes, and smoking [3]. Using genome-wide association studies, it was investigated that more than 164 genetic loci are involved in the pathogenesis of CHD. Interestingly, variations in the non-protein-coding regions of DNA disrupt the expression of CHD-associated genes.

Alteration in both the exonic and intronic regions of the genome leads to CHD. The non-coding parts of the genome encode short RNA molecules called microRNAs (miRNAs) [4]. Currently, 2654 mature miRNAs have been discovered in humans [5]. These RNA molecules are frequently linked to cell development, differentiation, and homeostasis and control the regulation of genes by repressing the translation or enhancing the destruction of complementary mRNA by binding to its 3′-UTR [6]. Dysregulated miRNAs are involved in the development and progression of multiple pathological conditions, including cardiovascular disease (CVD) [7]. Plasma *MIR145* may be a useful biomarker for coronary artery disease (CAD) and be able to predict CAD [8]. Among the Egyptian population, *MIR146A* rs2910164 and *MIR4513* rs2168518 may serve as a useful biomarker for CHD susceptibility [9]. The expression of *MIR484* is upregulated in acute coronary syndrome patients [10]. Elevated level of miR-126-3p has indicated linkage with cardiac pathologies [11]. *MIR21* is an important biomarker for atrial fibrosis [12]. *MIR21, MIR27B, MIR122, MIR125B, MIR146B, MIR147B*, and *MIR155* were less expressed while *MIR16* and *MIR92A* were highly expressed in premature CAD as compared to old age CAD patients [13]. In early onset CAD individuals, six miRNAs (miR340, -451, -454, -545, -615-5p, and 624) were upregulated and *MIR1280* was downregulated as compared to controls [14]. In comparison to healthy individuals, miR-208a-3p is substantially upregulated in CHD cases [15]. Patients with hypertension have higher levels of *MIR34A* in their peripheral blood [16]. The level of miR-483-3p in the serum was linked to the development of hypertension which is a major contributor to CHD and stroke [17]. A review reported that *MIR215, MIR487A*, and *MIR502* are diagnostic and prognostic biomarkers in stable CAD [18]. SNPs are the most common type of genetic variation in human genomes which are mostly located in non-coding regions [19]. MiR-SNPs change the levels of miRNA expression, which increases one’s vulnerability to a number of diseases [20]. SNPs alter phenotypes and are crucial in the progression of disease development [21]. It may have complicated impacts on miRNAs. Stability or processing may be impacted by these variations in immature miRNAs [22]. Such polymorphisms in the pri-miRNA promoters may affect the expression of mature miRNAs. Identification of the target gene may be impacted by SNPs in the seed region [23]. The seed region variant rs3746444 has been linked to an increased risk of CHD [24]. The rs3746444 genetic variation of *MIR499a* directly targets MTR (Methionine synthase) gene which may be closely linked with CHD risk in the Chinese cohort [25]. The genetic variant rs2910164 in pre-*MIR146A* is associated with a lower risk of acute coronary syndrome [26]. SNPs in *MIR146, MIR149, MIR196A2*, and *MIR499* genes have been associated with the pathophysiology of CAD [27].

Our study aims to assess the association of the respective variants in miRNAs (*MIR149, MIR499A, MIR196, MIR223*, and *MIR155*) with CHD pathogenesis.

## Results

### HWE of SNPs in miRNA Regions

150 CHD cases and 150 normal individuals were successfully genotyped to determine the association of SNPs in the relevant miRNAs. All cases and controls were in HWE at the studied SNPs located within miRNAs gene sequences (P<0.05).

### Allelic and Genotypic frequencies for rs3746444

For SNP, rs3746444 in *MIR499A*, the distribution of genotypes between cases and controls showed significant association using co-dominant model [χ2 =79.51; P = 0.0001], homozygous dominant model (GG vs AA+AG) [OR = 9.333 (5.180-16.82); P = 0.0001], heterozygous model (AG vs AA+GG) [OR = 0.1241 (0.065-0.234); P = 0.0001] and additive model [OR = 0.3440 (0.2468-0.4795); P = 0.0001] respectively. However, in the homozygous recessive model, the association of its genotypes was insignificant (AA vs GG+AG) [OR = 0.7130 (0.4471-1.137); P = 0.1921] as shown in Table. 02.

**Table. 01.** Presymptomatic data about the cases and controls.

| Categories | Gender | Mean Age (Year) | BMI (Kg/M<sup>2</sup>) | Mean B.P Systolic/Diastolic (mmHg) | DM | RBS (mg/dL) | LDL (mg/dL) | HDL (mg/dL) | TC (mg/dL) | TGs (mg/dL) |
| --- | --- | --- | --- | --- | --- | --- | --- | --- | --- | --- |
| **Cases (CHD)** | M= 106<br>F= 44 | 55.9 | 27.6 | 128/93.5 | Type-1<br>DM=136<br>Type-2<br>DM= 10<br>Healthy= 04 | 144.4 | 51.2 | 137.8 | 191.2 | 231.6 |
| **Controls** | M= 119<br>F= 31 | 58.8 | 26.2 | 124/92.4 | ----- | 101.9 | 85.5 | 101.7 | 173.4 | 131.3 |

blood sugar, LDL = Low density lipoprotein, HDL = High density lipoprotein, TC = Total cholesterol, TGs = Triglycerides

**Table. 02.** Statistical analysis for calculating the allele and genotype frequency of rs3746444, rs11614913, rs767649, rs1044165, and rs2292832 in CHD patients compared to healthy individuals using co-dominant, dominant, recessive, and additive models.

|  | Models | Genotype | Cases (n=150)<br>(% age) | Controls<br>(n=150)<br>(% age) | OR 95% CI | X<sup>2</sup>-<br>value | P-value |
| --- | --- | --- | --- | --- | --- | --- | --- |
| rs3746444 | Co-Dominant<br>Model | AA<br>AG<br>GG | 52(35%)<br>14(9%)<br>84(56%) | 64(43%)<br>68(45%)<br>18(12%) | - | 79.51 | 0.0001 |
|  | Homozygous<br>Dominant | GG<br>AA+AG | 84(56%)<br>66(44%) | 18(12%)<br>132(88%) | 9.333<br>(5.180-16.82) | - | 0.0001 |
|  | Homozygous<br>Recessive | AA<br>GG+AG | 52(35%)<br>98(65%) | 64(43%)<br>86(57%) | 0.7130<br>(0.4471-1.137) | - | 0.1921 |
|  | Heterozygous | AG<br>AA+GG | 14(9%)<br>136(91%) | 68(45%)<br>82(55%) | 0.1241<br>(0.065-0.234) | - | 0.0001 |
|  | Additive | A<br>G | 118(39%)<br>182(61%) | 196(65%)<br>104(35%) | 0.3440<br>(0.2468-0.4795) | - | 0.0001 |
| rs11614913 | Co-Dominant<br>Model | CC<br>CT<br>TT | 80(53%)<br>58(39%)<br>12(8%) | 56(37%)<br>58(39%)<br>36(24%) | - | 16.24 | 0.0003 |
|  | Homozygous<br>Dominant | CC<br>CT+TT | 80(53%)<br>70(47%) | 56(37%)<br>94(63%) | 1.918<br>(1.210-3.042) | - | 0.0075 |
|  | Homozygous<br>Recessive | TT<br>CT+CC | 12(8%)<br>138(92%) | 36(24%)<br>114(76%) | 0.2754<br>(0.1369-0.5540) | - | 0.0002 |
|  | Heterozygous | CT<br>CC+TT | 58(39%)<br>92(61%) | 58(39%)<br>92(61%) | 1<br>(0.6282-1.592) | - | 1 |
|  | Additive | C<br>T | 218(73%)<br>82(27%) | 170(57%)<br>130(43%) | 2.033<br>(1.445-2.861) | - | 0.0001 |
| rs767649 | Co-Dominant<br>Model | AA<br>AT<br>TT | 46(31%)<br>88(58%)<br>16(10%) | 8(5%)<br>36(24%)<br>106(71%) | - | 114.9 | 0.0001 |
|  | Homozygous<br>Dominant | AA<br>AT+TT | 46(31%)<br>104(69%) | 8(5%)<br>142(95%) | 7.851<br>(3.554-17.34) | - | 0.0001 |
|  | Homozygous<br>Recessive | TT<br>AT+AA | 16(11%)<br>134(89%) | 106(71%)<br>44(29%) | 0.04956<br>(0.026-0.092) | - | 0.0001 |
|  | Heterozygous | AT<br>AA+TT | 88(59%)<br>62(41%) | 36(24%)<br>114(76%) | 4.495<br>(2.737-7.382) | - | 0.0001 |
|  | Additive | A<br>T | 180(60%)<br>120(40%) | 52(17%)<br>248(83%) | 7.154<br>(4.902-10.44) | - | 0.0001 |
| rs1044165 | Co-Dominant<br>Model | AA<br>AG<br>GG | 4(3%)<br>6(4%)<br>140(93%) | 4(3%)<br>16(11%)<br>130(87%) | - | 4.91`6 | 0.0856 |
|  | Homozygous<br>Dominant | GG<br>AG+AA | 140(93%)<br>10(7%) | 130(87%)<br>20(13%) | 2.154<br>(0.9717-4.774) | - | 0.0819 |
|  | Homozygous<br>Recessive | AA<br>AG+GG | 4(3%)<br>146(97%) | 4(3%)<br>146(97%) | 1<br>(0.2453-4.076) | - | 1 |
|  | Heterozygous | AG<br>GG+AA | 6(4%)<br>144(96%) | 16(11%)<br>134(89%) | 0.3442<br>(0.1308-0.9055) | - | 0.0276 |
|  | Additive | G<br>A | 286(95%)<br>14(5%) | 276(92%)<br>24(8%) | 1.775<br>(0.9002-3.505) | - | 0.1305 |
| rs2292832 | Co-Dominant<br>Model | TT<br>CT<br>CC | 52(35%)<br>82(55%)<br>16(10%) | 56(37%)<br>66(44%)<br>28(19%) | - | 5.151 | 0.0761 |
|  | Homozygous<br>Dominant | TT<br>CT+CC | 52(35%)<br>98(65%) | 56(37%)<br>94(63%) | 0.8907<br>(0.5556-1.428) | - | 0.7183 |
|  | Homozygous<br>Recessive | CC<br>CT+TT | 16(11%)<br>134(89%) | 28(19%)<br>122(81%) | 0.5202<br>(0.2685-1.008) | - | 0.0717 |
|  | Heterozygous | CT<br>TT+CC | 82(55%)<br>68(45%) | 66(44%)<br>84(56%) | 1.535<br>(0.9733-2.420) | - | 0.0831 |
|  | Additive | C<br>T | 114(38%)<br>186(62%) | 122(41%)<br>178(59%) | 0.8942<br>(0.6443-1.241) | - | 0.5586 |

### Allelic and Genotypic frequencies for rs11614913

For SNP, rs11614913 in *MIR196*, the distribution of genotypes between cases and controls showed significant association using co-dominant model [χ2 =16.24; P = 0.0003], homozygous dominant model (CC vs CT+TT) [OR = 1.918 (1.210-3.042); P = 0.0075], homozygous recessive model (TT vs CT+CC) [OR = 0.2754 (0.1369-0.5540); P = 0.0002], and additive model [OR = 2.033 (1.445-2.861); P = 0.0001]. However, in the heterozygous model, the association of its genotypes was insignificant (CT vs CC+TT) [OR = 1 (0.6282-1.592); P = 1].

### Allelic and Genotypic frequencies for rs767649

For the rs767649 SNP of *MIR155*, it was noted that all the inheritance models showed significant association between the genotypes of cases and controls. The co-dominant model [χ2 =114.9; P = 0.0001], homozygous dominant model (AA vs AT+TT) [OR = 7.851 (3.554-17.34); P = 0.0001], homozygous recessive model (TT vs AT+AA) [OR = 0.04956 (0.026-0.092); P = 0.0001], heterozygous model (AT vs AA+TT) [OR = 4.495 (2.737-7.382); P = 0.0001], and additive model [OR = 7.154 (4.902-10.44); P = 0.0001] were statistically significant.

### Allelic and Genotypic frequencies for rs1044165

*MIR223* was studied for rs1044165 SNP and only the heterozygous inheritance model (AG vs GG+AA) [OR = 0.3442 (0.1308-0.9055); P = 0.0276] showed significant association (P □ 0.05) between the genotypes of cases and controls while the co-dominant model, homozygous dominant model, homozygous recessive model, and the additive model were insignificant (P □ 0.05).

### Allelic and Genotypic frequencies for rs2292832

The inheritance models i.e., co-dominant model, homozygous dominant model, homozygous recessive model, heterozygous model, and additive model showed the P value above 0.05 which represents statistically insignificant relationship between the genotypes of cases and controls for the SNP rs2292832 of *MIR149* gene as mentioned in the following table.

**The impact of variants on the Stem-loop structure of selected miRNAs**

The online tool RNAfold **(http://rna.tbi.univie.ac.at/cgi-bin/RNAWebSuite/RNAfold.cgi)** was used for calculating various crucial values for the investigated reference polymorphisms and their altered mutants of respective miRNAs as shown in Table. 03.

**Table. 03.**
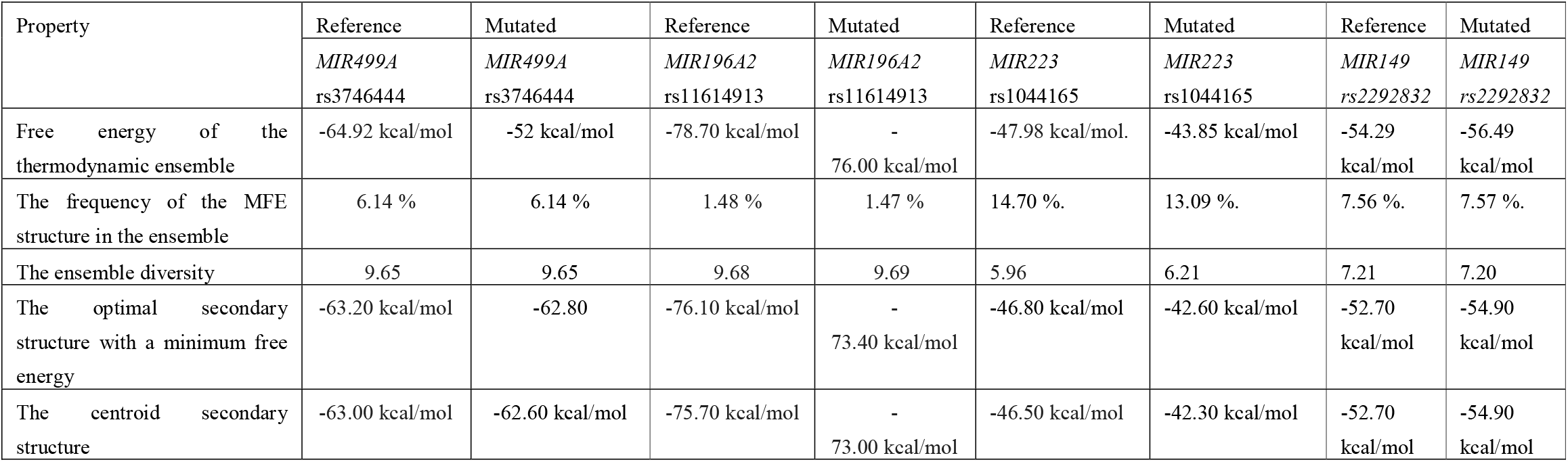
Information about the reference polymorphisms and their mutated variants.

It was observed that the relevant SNPs in *MIR499A, MIR196, MIR155*, and *MIR149* showed no effect on the secondary structure of these miRNAs while rs1044165 in *MIR223* changed its secondary structure by creating a visible stem loop. The current data shows that the variant can generate significant structural change in the precursor miRNA as shown in Figure. 01. This method allowed us to assess the effect of SNP on the secondary structure’s stability.

**Figure. 01.**
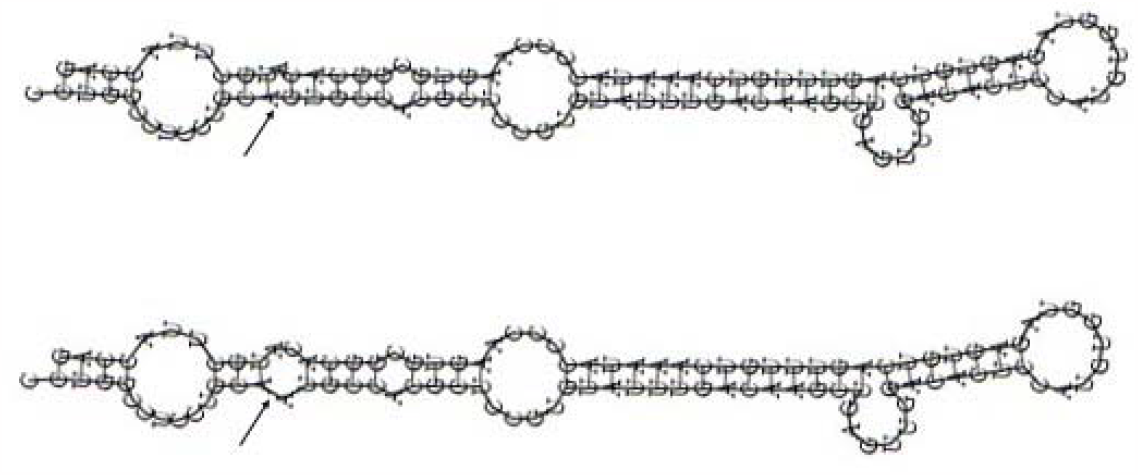
The upper structure shows the Stem-loop of reference *MIR223* while the lower shows creation of new loop due to variation in the *MIR223* gene.

## Discussion

Several human disorders have been connected to SNPs in protein-coding genes. Pre-miRNAs are formed by the transcription of miRNA genes to become mature miRNAs. Pre-miRNA SNPs may have a direct impact on miRNA maturation, expression, or binding to target mRNA that leads to dysregulation of target genes [28]. As a result, a functional SNP in a miRNA gene may modify the expression of downstream genes, affecting many signaling cascades. It has been demonstrated that such variations in these non-coding areas of the genome are linked to various diseases, including cancer and cardiovascular diseases [29-32]. The *MIR499A* is clustered with *MIR499B* and is located on the positive strand of Chromosome No.20 (genomic context from 34990376 to 34990497). The rs3746444 polymorphism is a functional variant of *MIR499A* which is formed of a nucleotide substitution from adenine (A) to guanine (G). A finding of the current study is in line with Labbaf et al where it was confirmed that the variant rs3746444 in *MIR499A* may contribute to the susceptibility of CAD [33]. Abdelghany et al. have confirmed that SNPs rs3746444 and rs2910164 in pre-miRNAs may increase the risk of ischemic stroke in the Egyptian population [34]. The *MIR196A2* gene is located on the positive strand of Chromosome No.12 (genomic context from 53991738-53991847) and its SNP rs11614913 is in the mature miRNA regions. It was investigated that the CC genotype in rs11614913 caused an increased expression of mature *MIR196A2* in the cardiac tissue samples of congenital heart disease in Chinese population [35]. A meta-analysis involving 16484 subjects by Liu et al revealed that increased risk is linked with *MIR146A* rs2910164, *MIR196A2* rs11614913, and *MIR499* rs3746444, but not with *MIR149* rs71428439 in patients with CHD [36].

In the present study, a population-based association of variants in different miRNA genes with CHD was investigated in the Pakistani population using TaqMan assay. Our findings suggest that *MIR499A, MIR196, MIR155*, and *MIR223* are strongly associated as risk factors for CHD susceptibility while the *MIR149* gene has no significant association with CHD pathogenesis within the Pakistani cohort.

As with any case-control study, one of the present study’s limitations is that it was impossible to rule out potential selection bias, which could have an impact on how the data are interpreted. The statistical power of our study may also be limited by the relatively small sample size. The role of these miRNA polymorphisms in CHD is still to be confirmed in larger-sample investigations in the Pakistani population.

## Materials and Methods

### Ethical approval and genomic DNA extraction

A proven history of CHD individuals was required for inclusion in this study, and individuals without such a history were not allowed to participate. Patients’ ages varied from 21 to 90, while controls’ ages ranged from 27 to 91 and they had no history of CHD. All participants in this study, including the healthy controls and patients, provided their informed consent. As demonstrated in Table.01., this study adhered to the principles of the Helsinki Declaration [37].

The Advanced Study and Research Board gave approval to this study in its 53^rd^ meeting held on January 08, 2020, at the University of Malakand, Pakistan. From CHD patients and healthy controls, 3-5 ml of whole blood was taken and stored in EDTA tubes at a 4□ cold temperature for further processing. The Phenol Chloroform method was employed to extract genomic DNA from the collected blood samples (healthy, n=150, and controls, n=150). The extracted DNA was dissolved in TE buffer and stored at a 4□ until further processing.

### TaqMan Assay

The miRbase (http://www.mirbase.org) contains information about the genetic background of 1917 precursors and 2654 mature human miRNAs. This database was used to obtain relevant data about the SNPs used in this study. Chromosomal locations and allele information for the miRNA variants rs2292832 in *MIR149*, rs3746444 in *MIR499A*, rs11614913 in *MIR196*, rs1044165 in *MIR223*, and rs767649 in *MIR155* were retrieved from miRbase [5]. TaqMan real-time PCR experiments were set up in 384-well plates comprising positive and negative samples and performed on an ABI Q6 system (Applied Biosystems). ABI-Prism 7900HT sequence detection devices were used to read and analyze the plates. The TaqMan assay was conducted double blinded to check the quality of the patients and healthy individuals.

### Statistical assessment

The Hardy-Weinberg Equilibrium (HWE) in the genotypes of the studied microRNA polymorphisms was investigated using the chi-square (χ2) test. The odds ratio (OR) and 95% confidence intervals (CI) were used to assess the relationship between the SNPs and the risk of CHD. This test determined the variance in genotype frequency distribution between patients and controls. To calculate the risk of CHD, various genetic inheritance models were applied. A low-risk allele was one whose frequency was lower in cases than in controls, while a high-risk allele was one whose frequency was higher in cases. The P-value for each genotype was calculated by comparing the homozygote and heterozygote genotype frequencies in patients and controls. Furthermore, the genotypic frequencies of the studied SNPs were calculated. A P-value of 0.05 or lower was regarded as statistically significant.

### *In-silico* Analysis

Alterations in Minimum Free Energy (MFE) between reference and mutated sequences in the stem-loop structure of miRNAs were calculated using RNAFold—ViennaRNA Package 2.0 [38] to predict the deleterious effects of SNPs in the studied miRNAs.

## Acknowledgments

The authors would like to express their gratitude to the patients and healthy controls who took part in this study.

## Footnotes

### Author contributions

Conceptualization: T.U.HAQ., A.A.SHAH.; Methodology: T.U.HAQ, Y.A, A.A.SHAH.; Validation: F. J.; Formal analysis: T.U.HAQ., A.A.SHAH.; Investigation: T.U.HAQ, S.A, S.U.R.; Writing - original draft: T.U.HAQ; Writing - review and editing: A.A.SHAH, Y.A, F. J.; Supervision: A.A.SHAH.; Funding acquisition: Y.C.

### Funding sources

The project was supported by Yangchao Chen, School of Biomedical Sciences, Chinese University of Hong Kong, NT, Hong Kong.

### Data Availability Statement

All the data is available in this manuscript.

### Conflict of interest

The authors declare that they have no conflicts of interest.

## Notes

### Competing Interest Statement

The authors have declared no competing interest.

### Author Declarations

This research synopsis was reviewed and recommended by the Departmental ethical committee (BEC). Furthermore, this research proposal was presented to the Graduate Study Committee (GSC). GSC is a departmental-level committee constituted by the vice-chancellor of the University of Malakand. The GSC evaluated this research synopsis for its suitability for ethical concerns and research design. GSC recommended this synopsis to The Advanced Studies and Research Board (ASRB) for further evaluation. ASRB is a university-level research evaluation body that checks all aspects of the submitted synopsis. The synopsis was again presented to ASRB by the first author. After extensive evaluation, the board granted approval to this research proposal in its 53rd meeting held on 08, January 2020, at the University of Malakand, Pakistan.

